# Immune responses to inactivated and vector-based vaccines in individuals previously infected with SARS-CoV-2

**DOI:** 10.1101/2022.01.03.22268704

**Authors:** Nungruthai Suntronwong, Ritthideach Yorsaeng, Chompoonut Auphimai, Thanunrat Thongmee, Preeyaporn Vichaiwattana, Sitthichai Kanokudom, Suvichada Assawakosri, Pornjarim Nilyanimit, Donchida Srimuan, Thaksaporn Thatsanatorn, Natthinee Sudhinaraset, Nasamon Wanlapakorn, Yong Poovorawan

**Affiliations:** Center of Excellence in Clinical Virology, Faculty of Medicine, Chulalongkorn University, Bangkok, 10330, Thailand; FRS(T), the Royal Society of Thailand, Sanam Sueapa, Dusit, Bangkok 10330, Thailand

## Abstract

Immunity wanes in individuals previously infected with SARS-CoV-2, and vaccinating those individuals may help reduce reinfection. Herein, reactogenicity and immunogenicity following vaccination with inactivated (CoronaVac) and vector-based (ChAdOx1-S, AZD1222) vaccines were examined in previously infected individuals. Immune response was also compared between short and long intervals between first date of detection and vaccination. Adverse events were mild but were higher with AZD1222 than with CoronaVac. Baseline IgG-specific antibodies and neutralizing activity were significantly higher with shorter than longer intervals. With a single-dose vaccine, IgG and IgA-specific binding antibodies, neutralizing activity, and total interferon-gamma response peaked at 14 days. Immune response was significantly higher in recovered individuals than in infection-naïve individuals. Antibody response was greater with longer than shorter intervals. AZD1222 induced higher antibody and T cell responses than those of CoronaVac. Thus, to achieve immunity, individuals with prior SARS-CoV-2 exposure may require only a single dose of AZD1222 or two doses of CoronaVac to achieve the immune response. These findings supported vaccine strategies in previously infected individuals.

## INTRODUCTION

Coronavirus disease 2019 (COVID-19) caused by the severe acute respiratory syndrome coronavirus-2 (SARS-CoV-2) continues to circulate globally and has been difficult to control even though COVID-19 vaccines are available^1^. Ongoing evolution and emergence of SARS-CoV-2 variants with significant mutations can make the virus more easily transmitted and less susceptible to neutralizing antibodies, slowing control of the pandemic^2,3^. In addition, immunity against SARS-CoV-2 wanes, even when induced through infection and vaccination, which has raised questions about long-term immunity^4,5^. A recent study indicated lower antibody titers against the spike protein of SARS-CoV-2 were associated with breakthrough infections^6^. Thus, vaccine strategies that promote robust protective immunity and maintain antibodies over time should be considered.

Natural infection with SARS-CoV-2 can elicit both humoral and T cell-mediated immune responses; however, length and severity of illness after SARS-CoV-2 infection were generally correlated with magnitude of immune response^7^. Infected individuals typically induce higher titers of neutralizing antibodies that target the viral spike glycoprotein (S)^8^. The SARS-CoV-2 spike protein is homo-trimeric and is cleaved into two subunits, S1 and S2^9^. The S1 subunit contains the N-terminal domain and a receptor-binding domain (RBD), which typically targets angiotensin-converting enzyme 2 (ACE2); both regions can potentially induce neutralizing activities. Simultaneously, the S2 subunit mediates viral membrane fusion during viral entry and is typically conserved across coronaviruses.

Although SARS-CoV-2 infection can produce relatively high levels of neutralizing antibodies that correlate with disease protection^10,11^, antibodies against SARS-CoV-2 appear to decline over time^12,13^. Antibody titers twelve months postinfection indicate that specific IgG against receptor binding sites decrease by 68.1% compared with that in the first month^14^. Despite a decline in antibody response, T cell-mediated immune response likely persists for several months and may persist longer than detectable antibodies in recovered patients^15,16^. Immunological memory to SARS-CoV-2 remains detectable for up to eight months following natural infection but gradually declines over a year^17,18^. However, rate of reinfection is reported as 1 per 100,000 persons, in at least one-year follow-ups^19^. Thus, vaccination should be considered for those with a history of previous infection in order to retain protective immunity.

Vaccination has been an effective tool in combatting COVID-19 pandemics^20^. Because of ongoing worldwide shortages of vaccines, whether a single dose or two doses are sufficient for recovered SARS-CoV-2 patients is under investigation. Vaccination of people who were exposed to SARS-CoV-2 with a single dose of mRNA vaccine can elicit a robust immune response that is higher than that with two doses of such a vaccine in individuals with no history of infection^21,22,23,24^. Similar results were found in a study of immunological memory after a single dose of a vector-based vaccine^25^. However, seropositive individuals may require two doses of CoronaVac to ensure induction of neutralizing antibodies^26^. Immunization of recovered individuals substantially increases humoral and cell-mediated immune responses against vaccine strains. It also improves neutralizing activities against SARS-CoV-2 variants with mutations in the spike protein, compared with uninfected individuals postvaccination^7^.

Although mRNA vaccine-induced immune response has been reported in individuals with past natural SARS-CoV-2 infection, few data are available for inactivated and vector-based vaccines in SARS-CoV-2 exposed individuals^25,26^. Moreover, additional data are needed to determine how the time interval between exposure and vaccination affects antibody dynamics and determines humoral and T cell responses after immunizing with inactivated or vector-based vaccine. Here, how individuals previously infected with SARS-CoV-2 develop an immune response when immunized with inactivated and vector-based vaccines was examined.

Immunogenicity was also compared in individuals with short and long intervals between infection and vaccination and in boosted responses between first and second doses. These results will offer insights into how immune response triggered by vaccination in individuals with prior infection should influence vaccine policies.

## RESULTS

### Study participants

There were 117 participants with a prior SARS-CoV-2 infection enrolled in the study. Two groups were designated according to the time interval between the first date of detection and vaccination: short interval (2 to 5 months) and long interval (13 to 15 months) (Fig. 1). Participants included 54 women (46.2%) and 63 men (53.8%). Ages ranged from 20 to 66 years, and mean age of participants immunized with CoronaVac (*n* = 58) was 40.3 (range: 20–57) and that of those who received AZD1222 (*n* = 59) was 42 (range: 25–66) (Table 1). The short interval group included 60 participants (51.3%), and the long interval group included 57 (48.7%). Baseline characteristics were similar between CoronaVac and AZD1222 recipients. Participants were classified into two groups per vaccine type according to the time interval between the first date of positive SARS-CoV-2 detection and vaccination (Supplementary Table 1). Mean age of CoronaVac recipients with short (*n* = 30) and long (*n* = 28) intervals was 37.6 and 43.1 years, respectively; whereas mean age of AZD1222 recipients with short (*n* = 30) and long (*n* = 29) intervals was 39.4 and 44.7 years, respectively. In both vaccine groups, mean age of previously infected participants with a long interval was higher than that of those with a short interval. Moreover, mean age of participants without prior infection was 42.6 and 47.6 years for CoronaVac and AZD1222, respectively.

**Table 1.** Demographic and characteristics of participants previously infected with SARS-CoV-2 enrolled in this study.

| Characteristics | CoronaVac<br>(n=58) | AZD1222<br>(n=59) | Overall<br>(n=117) |
| --- | --- | --- | --- |
| Female, n (%) | 23 (39.7 %) | 31 (52.5 %) | 54 (46.2 %) |
| Age, mean (range) | 40.3 (20- 57) | 42 (25- 66) | 41 (20- 66) |
| BMI (*kg m*<sup>-2</sup>) |  |  |  |
| BMI < 30 | 50 (86.2 %) | 54 (91.5%) | 104 (88.9 %) |
| BMI ≥ 30 | 8 (13.8 %) | 5 (8.5%) | 13 (11.1 %) |

| Comorbidities or Health conditions |  |  |  |
| --- | --- | --- | --- |
| DM | 2 (3.4 %) | - | 2 (1.7 %) |
| DM+HT | 1 (1.7 %) | 1 (1.7 %) | 2 (1.7 %) |
| HT | 1 (1.7 %) | 3 (5.1 %) | 4 (3.4 %) |
| HT+DM+CHD | 1 (1.7 %) | - | 1 (0.9 %) |
| Asymptomatic | 8 (13.8 %) | 8 (13.6 %) | 16 (13.7 %) |
| Symptomatic |  |  |  |
| w/o pneumonia | 34 (58.6 %) | 32 (54.2 %) | 66 (56.4 %) |
| w/ pneumonia | 16 (27.6 %) | 19 (32.2 %) | 35 (29.9 %) |
| Time interval between infection and vaccination |  |  |  |
| 2 to 5 months | 30 (51.7 %) | 30 (50.8 %) | 60 (51.3 %) |
| 13 to 15 months | 28 (48.3 %) | 29 (49.2 %) | 57 (48.7 %) |

Data presents as n (%), mean (range).
DM: Diabetes Mellitus, HT: Hypertension, CHD: Coronary heart disease

**Fig. 1:**
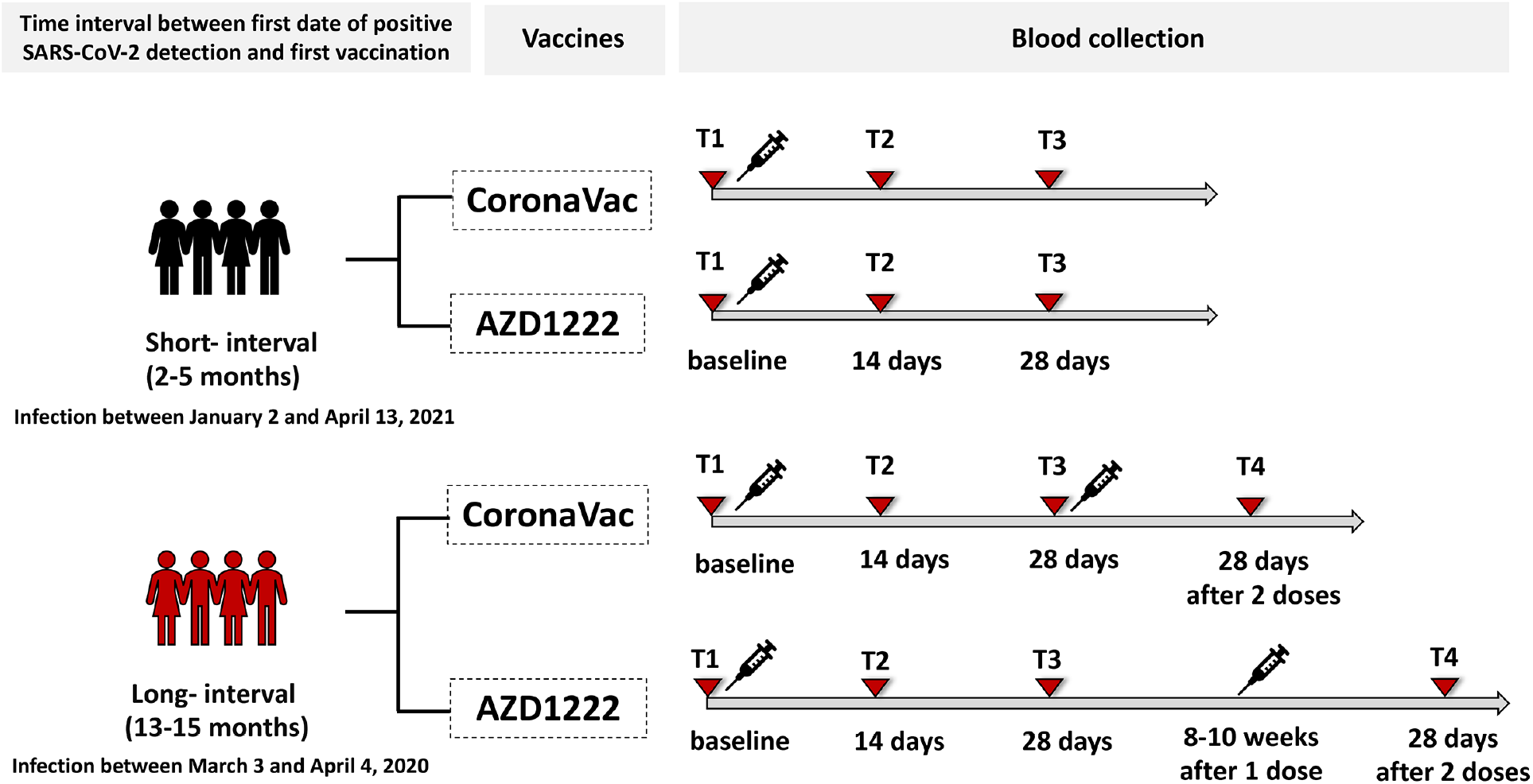
Schematic of vaccination timing and blood sampling. Previously infected individuals were classified into two groups according to the time interval between the first date of positive SAR-CoV-2 detection and vaccination: short interval (2–5 months) and long interval (13–15 months). Short-interval participants were infected with SARS-CoV-2 between January 2 and April 13, 2021, and long-interval participants were infected from March 3 to April 4, 2020. Those two groups each included one group that received CoronaVac and one group that received AZD1222. The short-interval group had a single-dose vaccination, and the long-interval group had a two-dose vaccination. Blood samples were collected at four different time points (0, 14, and 28 d after the first dose and one month after the second dose).

### Reactogenicity after vaccination

Reactogenicity analyses were based on adverse events (AEs) within 7 d after vaccination in 117 participants for the first dose and 50 participants for the second dose (Fig. 2). Local and systemic AEs were more frequent with the first dose than with the second dose. Most AEs were mild and moderate and were most frequent within 2 to 3 d after vaccination. In addition, AZD1222 recipients had a higher percentage of AEs than that of CoronaVac recipients (Supplementary Fig. 1). The most common AEs after receiving the first dose of AZD1222 were injection-site pain (84.8%), redness (23.7%), and swelling (22%) and systemic headache (79.7%), myalgia (81.4%), fever (17%), dizziness (17%), and fatigue (15.3%). After the first dose of the CoronaVac vaccine, local injection-site AEs included pain (55.2%), redness (5.2%), and swelling (3.5%), and systemic AEs included headache (34.5%), myalgia (34.5%), dizziness (5.2%), and fatigue (1.7%). However, serious AEs were not reported.

**Fig. 2:**
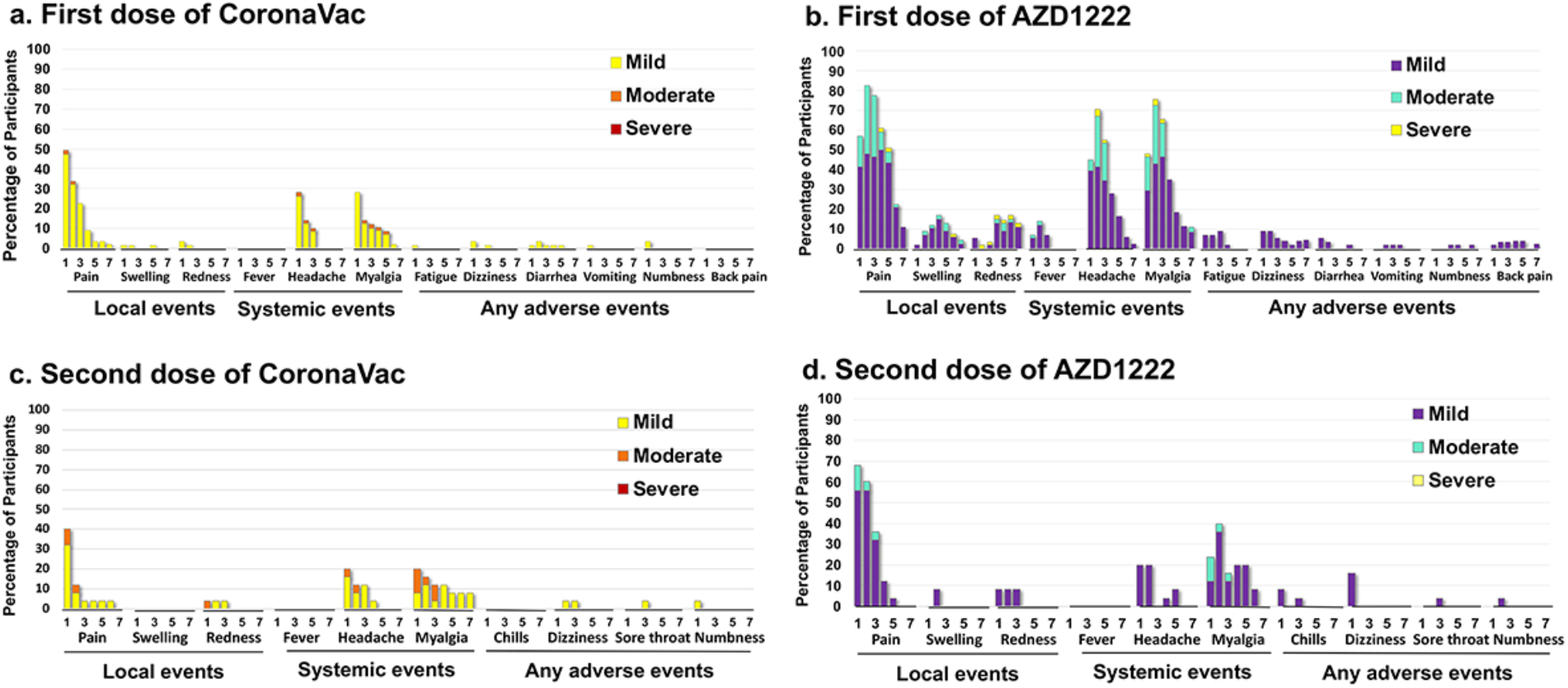
Solicited local and systemic adverse events within seven days (1, 3, 5, 7) after vaccination in participants previously infected with SARS-CoV-2. Bar graphs present adverse events following the first dose vaccination in 117 participants who received **a** CoronaVac (*n* = 58) and **b** AZD1222 (*n* = 59) and following the second dose of **c** CoronaVac (*n* = 50) and **d** AZD1222 (*n* = 50). Day 1 is the day of first and second dose vaccinations. Fever was classified as mild (38 °C to <38.5 °C), moderate (38.5 °C to <39 °C), and severe (≥39 °C). Swelling was graded as mild (<5 cm), moderate (5 cm to <10 cm), and severe (≥10 cm). Symptoms were graded as follows: mild, no limitation on normal activity; moderate, some limitation of daily activity; and severe, unable to perform or daily activity prevented.

### Antibody response following vaccination

Pre-existing immunity was compared between participants with short (2–5 months) and long (13–15 months) intervals. Before vaccination, 78.33% (47/60) and 98.33% (59/60) of participants with short intervals and 24.56% (14/57) and 92.98% (53/57) of participants with long intervals were seropositive for anti-N IgG and anti-S1 IgG, respectively, indicating exposure to SARS-CoV-2. As expected, although significantly higher IgG-specific SARS-CoV-2 antibodies were observed in short-interval participants than in their counterparts, IgG antibodies in participants with long intervals were detected above the cutoff level at baseline. This result indicated that IgG-specific antibodies, especially anti-RBD and anti-S IgG, persisted more than one year following natural infection. However, there was no difference in anti-S1 IgA between short and long intervals (Supplementary Fig. 2a-d).

Binding antibody response after vaccination was compared in participants with short and long intervals since infection and in those without prior infection. Levels of IgG antibodies specific to N and RBD and IgA-specific S1 were measured at different time points (Fig. 3). Following a single dose of CoronaVac, anti-N IgG seropositivity increased slightly but not significantly in participants with short intervals (Fig. 3a). By contrast, anti-N IgG increased significantly in those with long intervals after the first and second doses of the CoronaVac vaccine (*p* < 0.001). As expected, the level of anti-N IgG did not increase after AZD1222 vaccination.

**Fig. 3:**
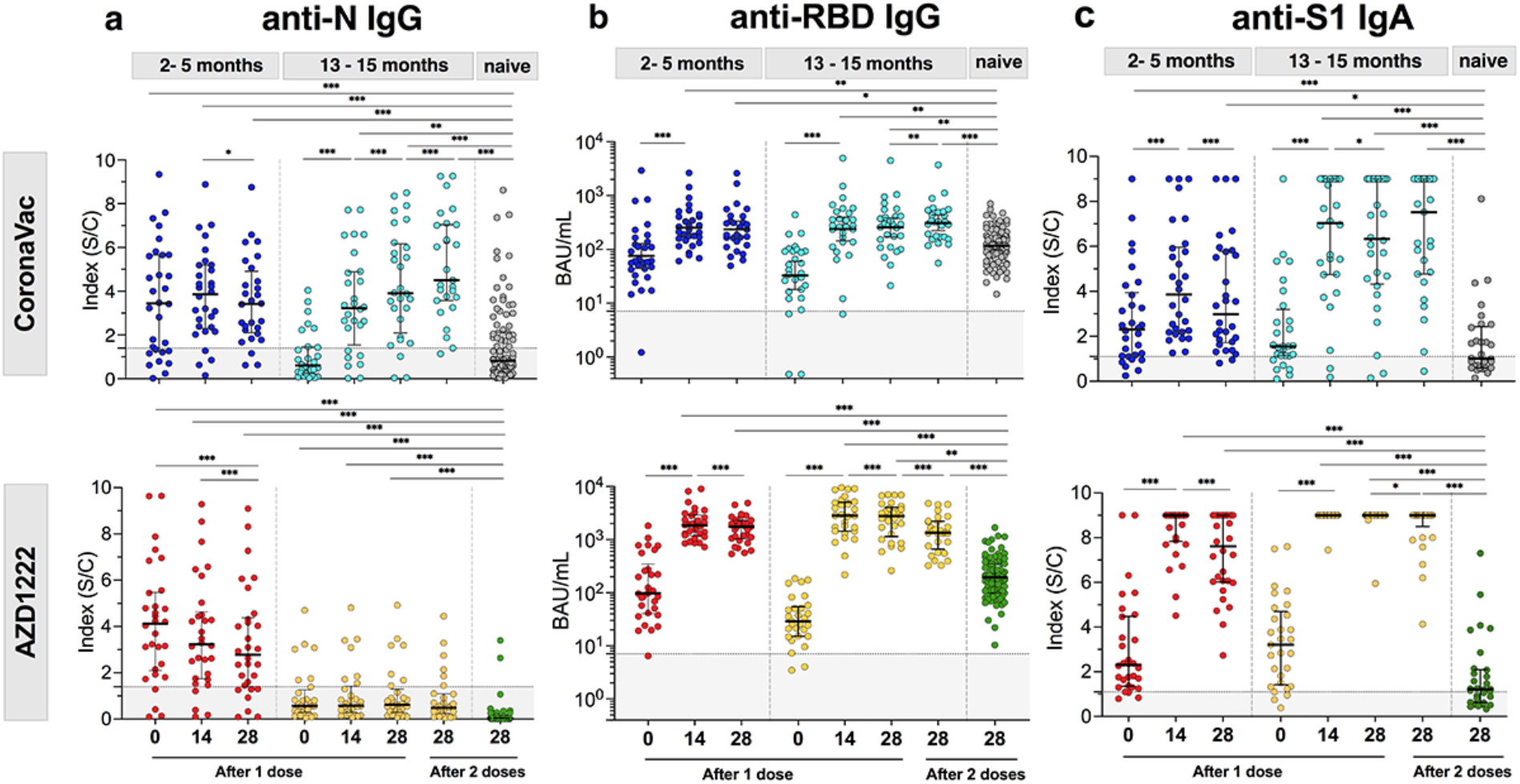
SARS-CoV-2-specific binding antibody response following CoronaVac and AZD1222 immunization in previously infected SARS-CoV-2 participants. Serum samples from CoronaVac and AZD1222-vaccinated individuals with prior infection (*n* = 117) were collected at 0, 14, and 28 d after the first dose and one month after the second dose to measure **a** anti-N IgG (optical density (OD) of sample divided by calibrator (Index S/C)), **b** anti-RBD IgG (binding antibody units per millilitre (BAU/mL)), and **c** anti-S1 IgA (Index S/C). Binding antibody level in previously infected individuals was compared between those with short (2–5 months, n=30 for CoronaVac, n=30 for AZD1222) and long (13–15 months, n=28 for CoronaVac and n=29 for AZD1222) intervals and with infection-naïve individuals (n=90 for CoronaVac and n=90 for AZD1222). Geometric mean titers with 95% confidence interval and median value with interquartile range are shown as horizontal bars. Dotted lines indicate cutoff values, and values under the cutoff are depicted in gray shaded areas. Statistics were calculated using Kruskal–Wallis tests with Dunns’ post hoc correction (\**p* < 0.05, \*\**p* < 0.01, and \*\*\**p* < 0.001).

The anti-RBD IgG titer in all vaccinated previously infected participants increased significantly and peaked at 14 d following a single dose of vaccine, compared with before vaccination (*p* < 0.001; Fig. 3b). At 28 d after single-dose vaccination with AZD1222, short and long-interval groups had levels of anti-RBD IgG that were 12.87-fold (1,549 binding antibody units per millilitre (BAU/mL) and 73.79-fold (2,258 BAU/mL) higher, respectively, than those of baseline. By contrast, after a single dose of CoronaVac vaccine, anti-RBD IgG levels were 3.12-fold (235 BAU/mL) and 7.79-fold (257 BAU/mL) higher in short and long-interval groups, respectively. Furthermore, anti-RBD IgG level increased significantly following the second dose of CoronaVac compared with that after the first dose (*p* < 0.01). By contrast, anti-RBD IgG level decreased significantly after the second dose of AZD1222, compared with the first dose (*p* < 0.001). However, a single dose of vaccine in recovered individuals led to higher anti-RBD IgG levels than those of infection-naïve individuals with complete two-dose vaccination (*p* < 0.01). A similar response was observed in levels of anti-S1 IgG (Supplementary Fig. 3).

In addition to measuring IgG, anti-S1 IgA levels were also tested, and anti-S1 IgA was detected in 84.61% (99/117) of participants before vaccination (Fig. 3c). At 28 d after the first dose of CoronaVac, anti-S1 IgA seropositivity in short and long-interval groups was 93.33% (28/30) and 92.85% (26/28), respectively. At 28 d after a single dose of AZD1222, 100% of previously infected participants were seropositive for anti-S1 IgA. Furthermore, with single-dose vaccination in recovered participants, anti-S1 IgA levels increased significantly compared with those in uninfected individuals postvaccination (*p* < 0.001).

### Neutralizing activities against SARS-CoV-2 wild type and variants

In addition to the binding antibody response, antibody function was further determined using a surrogate virus neutralization test (Fig. 4). First, in serum samples collected at different time points, presence of neutralizing antibodies against wild-type SARS-CoV-2 was determined using a NeutraLISA assay based on blockage of the ACE2–RBD protein–protein interaction. Previously infected individuals had a strong response following a single dose of vaccine, and 87.93% (51/58) of those with CoronaVac and 100% (59/59) of those with AZD1222 developed neutralizing activities. Those levels of neutralizing activity were significantly higher than those in infection-naïve individuals (63.33% (57/90) and 92.22% (83/90) for two doses of CoronaVac and AZD1222, respectively, *p* < 0.001) (Fig. 4a, b). In participants with prior infection, percentages of neutralizing activities in CoronaVac recipients (84.6% and 94.8% for short and long intervals, respectively) were significantly lower than those in AZD1222 recipients (99.4% and 99.5% for short and long intervals, respectively) (*p* < 0.001). Neutralizing capacity increased after the second dose of vaccine, reaching 95.4% (interquartile range (IQR) : 77.6% to 98%) for CoronaVac and 99.4% (IQR: 98.7% to 99.5%) for AZD1222.

**Fig. 4:**
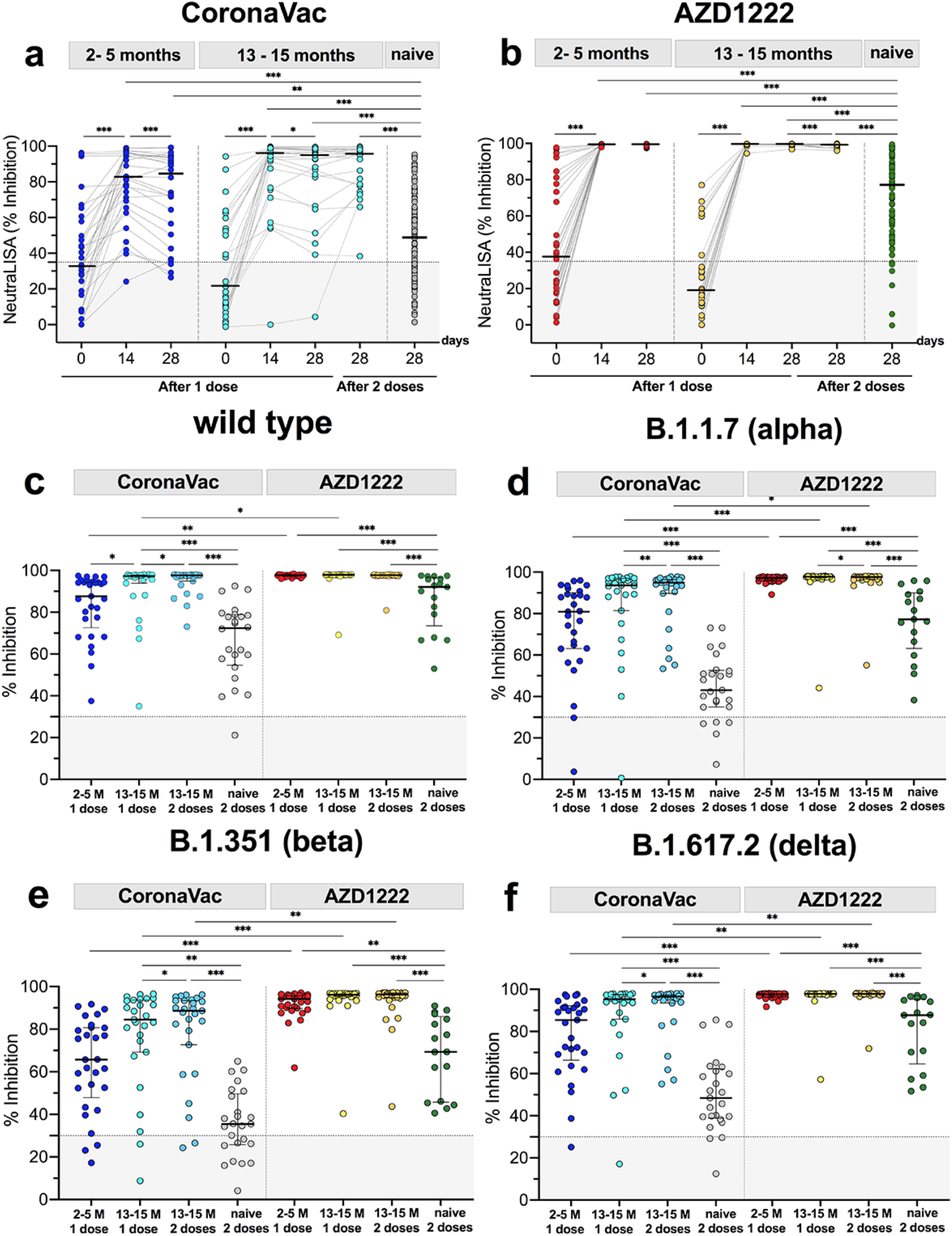
Neutralizing activity against SARS-CoV-2 wild type and variants. Serum samples from **a** CoronaVac and **b** AZD1222-vaccinated participants were measured for neutralizing activity on days 0, 14, and 28 after the first dose of vaccine and one month after second doses using a surrogate viral neutralization test (NeutraLISA, Euroimmun). Serum samples on day 28 after first and second doses of vaccinations were measured for neutralizing activity against **c** the wild type and **d** B.1.17 (alpha), **e** B.1.352 (beta), and **f** B.1.617.2 (delta) variants using cPass^™^ neutralization kits. Median values with interquartile ranges are depicted as horizontal bars. Dotted lines indicate cutoff values, and values under the cutoff are depicted in gray shaded areas. Serum-neutralizing activities in vaccinated previously infected individuals with 2–5-month (n=30 for CoronaVac and n=30 for AZD1222) and 13–15-month intervals (n=27 for CoronaVac and n=29 for AZD1222) were compared with those in serum samples from infection-naïve individuals after two doses of CoronaVac (n=90) or AZD1222 (n=89) vaccine. Statistics were calculated using Kruskal–Wallis tests with Dunns’ post hoc correction (\**p* < 0.05, \*\**p* < 0.01, and \*\*\**p* < 0.001).

Neutralizing potential of sera collected one month after first and second doses of vaccines against SARS-CoV-2 wild type and variants was measured using cPass^™^ ELISA assays (Fig. 4c–f). A single dose of vaccine was very influential in broadly inducing antibodies to SARS-CoV-2 variants in almost all participants with prior infection, and antibody function was significantly higher than that in infection-naïve individuals (*p* < 0.001). All participants who experienced infection had detectable neutralizing activity against the wild type, based on the cPass^™^ assay (Fig. 4c). Nevertheless, 27/29 (93.1%) and 26/27 (96.3%) of individuals with short and long intervals, respectively, showed positive neutralization against B.1.1.7 following a single dose of CoronaVac (Fig. 4d). All participants vaccinated with AZD1222 had strong neutralizing activity, with blockage of ACE2–RBD exceeding 97%. As expected, there was a decrease in SARS-CoV-2 neutralizing activity against the B.1.351 variant (Fig. 4e).

Notably, more than 96% of individuals with previous infection had a robust increase in neutralizing antibodies against B.1.617.2 after a single dose of CoronaVac, and the percentage reached 100% for the AZD1222 vaccine (Fig. 4f). Overall, neutralizing activity increased significantly after two doses of the CoronaVac vaccine. However, there was no significant increase in neutralizing activity against SARS-CoV-2 variants following the second dose of AZD1222, except against B.1.1.7. Furthermore, participants with long intervals had higher levels of neutralizing activity than those with short intervals. These findings indicated that neutralizing activity after vaccination was greater in participants with longer intervals than in those with shorter intervals.

### Total interferon-gamma response following immunization

Total IFN-γ responses stimulated by CD4+ epitopes derived from the S1 subunit (RBD) (Ag1) and by CD4+ and CD8+ epitopes derived from S1 and S2 subunits of the spike protein (Ag2) were investigated. Subtracted interferon-gamma responses (the level of interferon gamma from Ag1 or Ag2 minus those level from negative control (Nil)) increased significantly 14 d after the first dose of both vaccines, compared with the baseline (Fig. 5). Number of seropositives was lower in CoronaVac recipients (73.33% (22/30) for short intervals; 74.07% (20/27) for long intervals) (Fig. 5a, b) than in AZD1222 recipients (93.33% (28/30) for short intervals; 93.1% (27/29) for long intervals) (Fig. 5c, d). In addition, immunization in the short-interval group sustained the interferon-gamma level at day 28 compared with that at day 14. When total interferon-gamma response was compared between first and second doses, CoronaVac recipients had a slight increase in interferon-gamma response for Ag2 after two doses of the vaccine (Fig. 5e). By contrast, level of interferon-gamma response after two doses of AZD1222 decreased significantly (Fig. 5f). These results suggest that a single dose of AZD1222 and two doses of CoronaVac are sufficient to induce total interferon-gamma response in individuals previously infected with SARS-CoV-2.

**Fig. 5:**
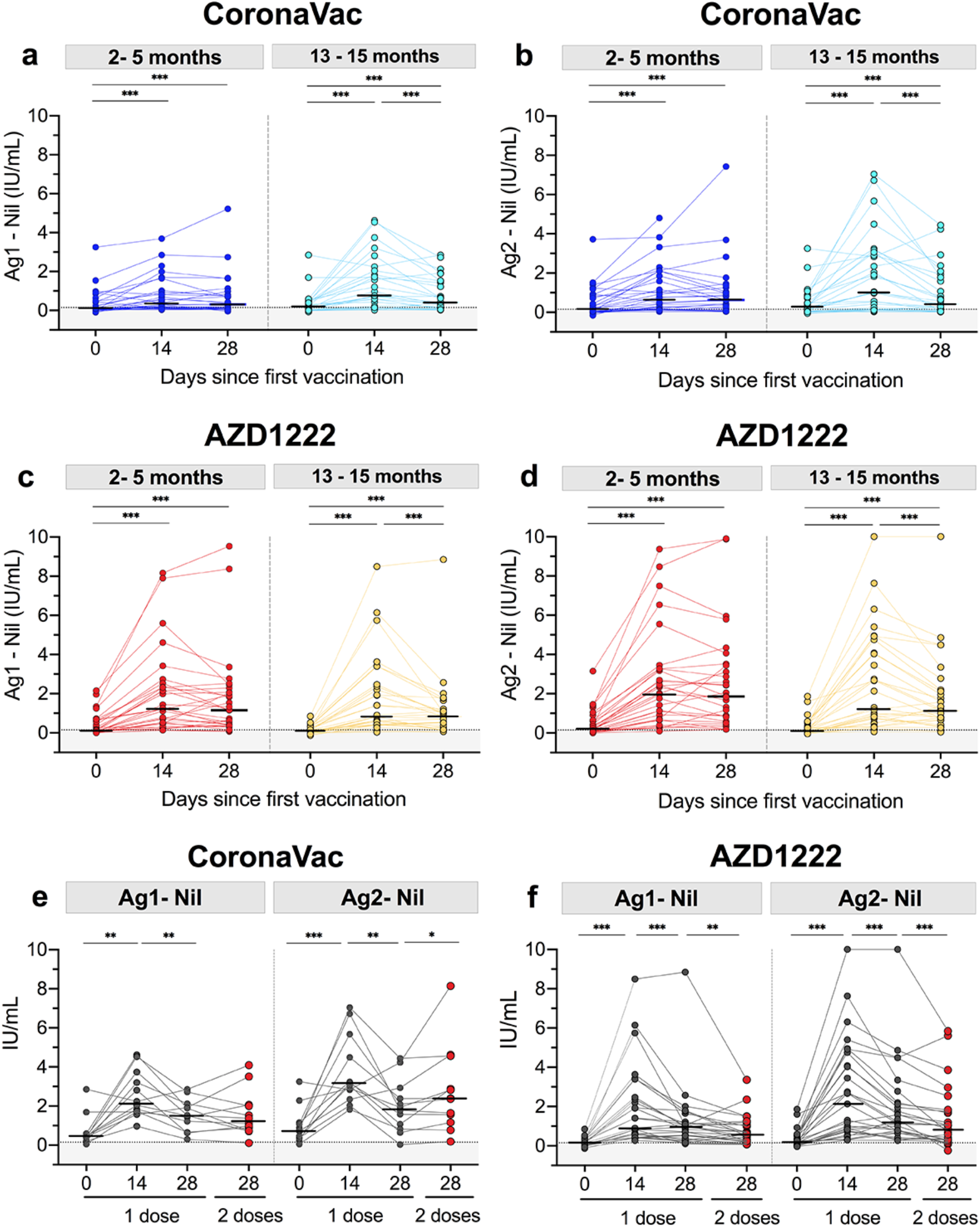
Comparison of total interferon-gamma-releasing T-cell responses to SARS-CoV-2 antigens. Serum samples from participants that received the CoronaVac (n=30 for 2-5 months and n=27 for 13-15 months) vaccine on days 0, 14, and 28 after the first dose were stimulated by **a** Ag1, which is a CD4+ epitope derived from RBD, minus negative control (Nil), and **b** Ag2, which is CD4+ and CD8+ epitopes derived from S1 and S2 subunits, minus negative control (Nil). Total interferon-gamma response of those with the AZD1222 (n=30 for 2-5 months and n=29 for 13-15 months) vaccine to SARS-CoV-2 **c** Ag1 and **d** Ag2 after the first dose at different time points. Interferon-gamma responses above cutoff values (0.15 international units per millilitre (IU/mL)) were detected in those with **e** CoronaVac (*n* = 12) and **f** AZD1222 (*n* = 27) vaccines after the second dose. Horizontal bars indicate the median. Two-tailed pair-matched comparisons were performed using Wilcoxon signed-rank tests. Statistical significance is indicated as follows: \**p* < 0.05, \*\**p* < 0.01, and \*\*\**p* < 0.001.

## DISCUSSION

Vaccination of individuals with prior SARS-CoV-2 infection by either inactivated or vector-based vaccine resulted in potent immune responses with titers exceeding those of unexposed individuals with two doses of vaccine. Consistent with recent studies, a single dose of vector-based vaccine or two doses of inactivated vaccine induced a robust immune response in seropositive individuals^25,26^. Notably, the second dose of AZD1222 did not provide additional boosting effects compared with those of the first dose in recovered patients. Previous reports indicate that improvement in vaccine-induced immune response might be related to extended vaccine dosing intervals^27,28,29^. By contrast, antibody level was higher after the second dose of CoronaVac than after the first dose. These results suggest that receiving either a single dose of vector-based vaccine or two doses of inactivated vaccine can recall adequate memory for both antibody and T-cell responses in previously infected individuals.

Overall, antibody titers were boosted significantly at 14 d after a single dose of vaccine, indicating a substantial B-cell memory response^30^. A similar result was found in seropositive individuals following a single dose of mRNA vaccine, with antibody titers gradually increasing over seven days and reaching a peak at day 14 after immunization^24,31^. CoronaVac contained the SARS-CoV-2 nucleocapsid protein, whereas AZD1222 did not. Therefore, only immunization with the inactivated vaccine significantly induced anti-N IgG. A long-term study indicates that anti-N antibodies can be detected for several months after natural infection^32^. Additionally, anti-N antibodies may protect against SARS-CoV-2 disease by promoting T-cell immunity^33^.

After boosting, anti-RBD and S1-specific IgG levels were significantly higher with AZD1222 than with CoronaVac. Although there was greater decay of anti-S1 and anti-RBD IgG after AZD1222 vaccination, a relatively high level of neutralizing activity against the wild type and SARS-CoV-2 variants persisted, indicating vaccination following infection could sustain prolonged protection^34^. Level of an immune correlate of protection against SARS-CoV-2 disease has not been established, but reduction in viral replication in nonhuman primates most strongly correlates with levels of anti-S and neutralizing antibodies after viral challenge^35^. In addition, protection against symptomatic B.1.1.7 variant infection is associated with 506 BAU/mL of anti-RBD IgG^36^. In AZD1222 recipients, but not in CoronaVac recipients, levels of anti-RBD IgG were higher than 506 BAU/mL, particularly in those with longer intervals. This result suggests that a single dose of AZD1222 can potentially protect against reinfection with SARS-CoV-2.

In addition to significantly elevated IgG levels, anti-S1 IgA was significantly boosted in previously infected individuals following vaccination with either inactivated or vector-based vaccine. Similar results are observed in seropositive individuals following a single dose of mRNA^29,37^. Although neutralizing activity of serum SARS-CoV-2-specific IgA is twofold lower than that of IgG equivalents^38,39^, IgA has been detected earlier and is more closely associated with neutralization than IgG in the first weeks after onset of symptoms. Such results suggest that serum IgA may reach the airway through transduction^40^. In addition, mucosal IgA has more potent neutralization activity than that of serum IgA and more rapid mobilization in response to SARS-CoV-2 infection. Unfortunately, the boosting effect of inactivated or vector-based vaccine on mucosal IgA response was not investigated in this study.

A single dose of vaccine in individuals who previously encountered SARS-CoV-2 infection led to an increase in neutralization potency that inhibited SARS-CoV-2 variants, including B.1.17, B.1351, and B.617.2. Consistent with a previous study, immunization in people with prior infection increased the range of neutralizing activity against SARS-CoV-2 variants^41^. Hybrid immunity suggests that those exposed to SARS-CoV-2 infection with subsequent COVID-19 vaccination can have a larger-than-expected, potent immune response^42^. Natural immunity combined with vaccine-induced immunity can provide broad neutralizing potency that is 25 and 100 times higher than that provided by vaccination and infection alone, respectively^43^. Increasing the range of neutralizing activity to protect against SARS-COV-2 variants after vaccination in previously infected individuals might be driven in part by immunological memory of B and T cells^44^.

Although protection against SARS-CoV-2 reinfection is correlated with level of antibodies, T-cell response may also contribute^36^. Notably, in this study, memory of T cell-related interferon-gamma release was awakened quickly after vaccination in recovered patients. According to a previous study^45^, differences in magnitude and quantity of T-cell response indicate that vaccination in previously infected individuals leads to a more robust spike-specific CD4 T-cell response and a higher level of CD8+ T-cell response than those in vaccinated naïve individuals. Total interferon-gamma-secreting T-cell response as evaluated by SARS-CoV-2 peaks at 11 to 14 d and gradually declines at 28 to 32 d after vaccination^46^. However, the SARS-CoV-2-specific T-cell response is maintained for several months after natural infection and vaccination^16^. Additionally, a recent study highlighted the potential for variants to escape from neutralizing humoral immunity but not from cell-mediated immunity^47^. Moreover, in a previous study, mutations in spike epitopes did not impair T-cell responses, and escaping neutralizing antibodies indicated that T cells might play a significant role in broadly protecting against SARS-CoV-2 variants^48^.

With both vaccines, a long interval between infection and vaccination led to a better immune response than that of a short interval. These findings are consistent with those of a study that found an extended gap between infection and vaccination increased peak antibody response after receiving a single dose of mRNA vaccine^29^. Furthermore, people with longer intervals had higher broadly neutralizing activity against SARS-CoV-2 variants than those with shorter intervals, indicating that RBD-specific memory B cells are relatively sustained for more than six months^41^.

The AE symptoms in previously infected individuals after vaccination were similar to those in individuals without prior infection. Therefore, pre-existing immunity after natural infection may not affect AE symptoms^49,50,51^. Additionally, AEs were more frequent after vaccination with the vector-based vaccine than with the inactivated vaccine, suggesting that different types of vaccines cause different types of local and systemic reactogenicity. Additionally, serious AEs were not observed after immunizing previously infected individuals with either an inactivated or a vector-based vaccine.

There were some limitations to the study. First, neutralizing titers were not examined. Laboratory testing was limited, because titers of anti-S1 IgA, neutralizing activity, and total interferon-gamma responses exceeded upper detection limits. Thus, exact levels of immune response after boosting could not be summarized. Moreover, other SARS-CoV-2-particular proteins, such as matrix and nucleocapsid proteins, which induce total interferon-gamma release^52^, did not stimulate SARS-CoV-2-specific T cells. Last, small sample size was a limitation. Participants were not randomly assigned to receive different types of vaccine, and vaccine groups were assigned on the basis of convenience.

Further investigation is warranted to assess the durability of antibody and T-cell responses after vaccination of people with previous SARS-CoV-2 infection, as well as the interplay between natural and vaccine-induced immunity. Notably, some vaccinated participants in this study failed to produce a booster response, particularly a T-cell response; thus, further studies in this population are needed. Longitudinal studies are required to determine persistence of humoral and T-cell responses after vaccination in previously infected individuals. Additionally, whether immunization of those with prior infection affects the level of mucosal IgA should be further investigated.

In conclusion, previously infected SARS-CoV-2 individuals developed a more robust immune response after AZD1222 or CoronaVac vaccination than that in naïve individuals. Notably, vaccination with AZD1222 produced higher antibody levels, neutralizing activity, and T-cell responses than those with the CoronaVac vaccine. In addition, a longer interval between infection and first vaccination improves the immune response more than that of a shorter one. The results suggest that a single dose of AZD1222 or two doses CoronaVac can boost antibody and T-cell responses in individuals with prior infection, which is a conclusion that can be paramount in facilitating vaccine policies.

## Methods

### Study design and participants

One hundred and seventeen recovered COVID-19 patients who were healthy adults aged ≥18 years and previously infected with SARS-CoV-2 (defined as anti-nucleocapsid positivity (IgG) or a history of positive SARS-CoV-2 detection) were enrolled with written consent. According to the duration between the first date of positive SARS-CoV-2 detection and vaccination, there were two groups of participants: the short-interval group with 2 to 5 months between dates (infection between January 2 and April 13, 2021), which included 60 participants, and the long-interval group with 13 to 15 months between dates (infection between March 3 and April 4, 2020), which included 57 participants (Fig. 1). Participants in each group were assigned to CoronaVac or AZD1222 vaccine on the basis of convenience. Participants were first vaccinated on May 14, 2021, and the last blood sample was collected on August 13, 2021. Furthermore, the short-interval groups received a single dose and the long-interval groups received two doses of the vaccines. Participants received a second dose of CoronaVac 28 d after the first dose and that of AZD1222 8 to 10 weeks after priming. For comparison as reference groups, data on 180 infection-naïve individuals who had been vaccinated with two doses of CoronaVac (*n* = 90) or AZD1222 (*n* = 90) were obtained from a previous study^50^. All participants have written consent.

Reactogenicity data were collected as self-reported AEs 7 d after the first and second doses of vaccines. According to the US Food and Drug Administration, solicited local and systemic AEs were classified as mild (no limitation on normal activity), moderate (some limitation of daily activity), and severe (unable to perform or prevented daily activity)^53^. Peripheral blood was collected before vaccination (day 0),14 and 28 d after the first dose, and one month after the second dose (Fig. 1). The study protocol was performed under the Declaration of Helsinki and Good Clinical Practice principles. Approvals were received from the Research Ethics Committee of the Faculty of Medicine, Chulalongkorn University (IRB numbers 192/64 and 281/64). This study has been registered with the Thai Clinical Trials Registry (TCTR20210319003 and TCTR20210520004). Informed consent was obtained before participant enrollment.

### Study vaccine

CoronaVac (0.5 mL dose) is an inactivated virus vaccine generated by growing the SARS-CoV-2 virus (CZ02 strain) in African green monkey kidney cells (Vero Cell), followed by inactivation with β-propiolactone and formaldehyde and adsorption with aluminum hydroxide^54^. The ChAdOx1-vectored vaccine (0.5 mL dose) is a recombinant, replication-deficient chimpanzee adenovirus-vectored vaccine, expressing the SARS-CoV-2 spike surface glycoprotein^49^.

### Serological testing

In all sera samples, anti-SARS-CoV-2 IgG and IgA immunoglobulin, including anti-nucleocapsid (N) protein IgG, anti-spike1-specific IgA, and anti-spike/receptor-binding domain (RBD)-specific IgG antibodies, were determined. According to the manufacturer’s instructions, anti-N IgG was determined using a SARS-CoV-2 IgG assay (Abbott Diagnostics, Abbott Park, IL, USA). Antibody amounts are presented as the IgG ratio (optical density (OD) divided by calibrator), with values ≥1.4 scored positive. The anti-SARS-CoV-2 S1 spike domain IgA was detected using enzyme-linked immunosorbent assay (ELISA) (Euroimmun, Lübeck, Germany), and results are expressed as the IgA ratio (≥1.1 positive).

The IgG antibodies against SARS-CoV-2 S1/RBD were quantitatively measured using CE marked SARS-CoV-2 Quant IgG II (Abbott Diagnostics), and antibody level is expressed as BAU/mL, with values ≥7.1 BAU/mL defined as positive. In addition, anti-S1 IgG was measured using LIASON SARS-CoV-2 Trimeric S IgG (DiaSorin, S.p.A, Saluggia, Italy), with results reported in BAU/mL and with a cutoff ≥33.8 BAU/mL defined as positive.

### Surrogate virus neutralization tests for SARS-CoV-2 wild type and variants

Two commercial kits were used for surrogate virus neutralization tests (sVNT) to determine neutralizing activity against SARS-CoV-2. The assay was based on antibodies-mediated blockage of angiotensin-converting enzyme 2 (ACE2)–RBD protein–protein interaction. In all serum samples, neutralizing activity against the wild type was determined by using enzyme-linked immunosorbent assay (ELISA)-based sVNTs according to the manufacturer’s instructions (Euroimmun). Results are shown as % inhibition (IH), and the positive cutoff was defined as ≥35%.

In addition, samples collected one month after the first and second doses of vaccines were tested with sVNTs against SARS-CoV-2 wild type and variants using a cPass^™^ SAR-CoV-2 neutralizing antibody detection kit according to the manufacturer’s instructions (GenScript, Jiangsu, China). In this assay, sera samples and positive and negative controls were diluted 1:10 with sample dilution buffer and then pre-incubated for 30 min at 37 °C with horseradish peroxidase-conjugated recombinant SARS-CoV-2 RBD protein of the wild type and B.1.1.7 (Alpha; N501Y), B.1.351 (Beta; K417N, E484K, and N501Y), and B.1.617.2 (Delta; L452R and T478K) variants. After pre-incubation, 100 µL of sample mixture was added onto human-angiotensin-converting enzyme 2 protein-coated ELISA plates for 15 min at 37 °C. ELISA plates were then washed three times with washing buffer, and 100 µL of 3,3′,5,5′-tetramethylbenzidine solution was added to develop a colorimetric signal. Plates were incubated in the dark for 15 min at 20 °C to 25 °C. Then, 50 µL of stop solution was added to quench the reaction, and the absorbance was immediately read at 450 nm. Percent inhibition for an individual sample was calculated as inhibition (%) = (1 - OD value of sample/average OD of negative control) × 100. Values above 30% indicated the presence of neutralizing antibodies.

### Quantification of interferon-gamma response

The SARS-CoV-2-specific T-cell response was evaluated by conducting a whole-blood Interferon-Gamma Release Assay according to the manufacturer’s instructions (QuantiFERON, Qiagen, Hilden, Germany). In this assay, whole heparinized blood was collected and then transferred to blood collection tubes coated with SARS-CoV-2-specific antigen peptides to stimulate cell-mediated immunity. The blood collection tubes consisted of two antigen tubes and positive and negative controls. Antigen tubes were coated with CD4+ epitopes derived from the S1 subunit (RBD) (Ag1) and CD4+ and CD8+ epitopes derived from S1 and S2 subunits of the spike protein (Ag2). After 1 mL of whole blood was added, QFN collection tubes were incubated for 24 h at 37 °C and then centrifuged to collect the plasma. The interferon-gamma release from stimulated samples was detected using ELISA according to the manufacturer’s guidelines (Qiagen). Following ELISA, interferon-gamma concentration was quantified based on the eight-point standard (0.125 to 8 IU/mL) and calculated as IU/mL by QuantiFERON RD (v5.03) software. The lower detection limit was 0.065 IU/mL, and IFN-g values ≥10 IU/mL were defined as 10 IU/mL. Results of IFN-g from Ag1 and Ag2 were subtracted from unstimulated controls (Nil) to represent the interferon-gamma response related to the SARS-CoV-2-specific T-cell stimulation. After subtraction, values ≥0.15 IU/mL indicated an elevated interferon-gamma response.

### Statistical analyses

Baseline characteristics of participants are presented as numbers or percentages, and categorical variables were computed using Chi-squared or Fisher’s exact tests. Age and time since infection are reported as the mean and SD and were compared with unpaired *t*-tests. Outcomes are reported as geometric mean titer (GMT) with 95% confidence intervals (95% CI) and median with interquartile range (IQR). Comparison of immune responses between groups was performed using Kruskal–Wallis tests with Dunns’ post hoc correction, and comparison within groups (different time points) was performed using Wilcoxon signed-rank tests. All statistical analyses were conducted using GraphPad Prism v9.0 (GraphPad, San Diego, CA) and SPSS v23.0 (IBM Corp, Armonk, NY). A *p*-value <0.05 was considered statistically significant.

## Data availability

All data are available in the main text. Additional information can be requested through the corresponding author.

## Data Availability

All data produced in the present work are contained in the manuscript.

## Acknowledgements

The authors are grateful to the participants in this study. We would like to thank all the staffs of the Center of Excellence in Clinical Virology for helping and supporting in this project. This work was supported by the National Research Council of Thailand (NRCT), Health Systems Research Institute (HSRI), the Center of Excellence in Clinical Virology of Chulalongkorn University, King Chulalongkorn Memorial Hospital, MK Restaurant Group, the Department of Disease Control (DDC), the Department of Medical Services Ministry of Public Health (MoPH), the Bangkok Metropolitan Administration (BMA) and National Blood Centre. Nungruthai Suntronwong was supported by the second Century Fund (C2F) fellowship of Chulalongkorn University.

## Author Contributions

Conceptualization, N. Suntronwong, N. Sudhinaraset, P.N., N.W. and Y.P.; data collection, N. Suntronwong, R.Y., D.S. and T. Thatsanatorn; formal analysis, N. Suntronwong, N.W. and Y.P.; methodology, N. Suntronwong, C.A., T. Thongmee, P.V., S.K. and S.A., and ; project administration, Y.P.; writing-original draft, N. Suntronwong and Y.P.; writing-review and editing, N. Suntronwong, N. Sudhinaraset, N.W. and Y.P.; All authors have read and agreed to the published version of the manuscript.

## Competing interests

All authors declared no conflict of interest

## Notes

### Competing Interest Statement

The authors have declared no competing interest.

### Clinical Trial

TCTR20210319003 
TCTR20210520004

### Author Declarations

The study protocol was performed under the Declaration of Helsinki and Good Clinical Practice principles. Approvals were received from the Research Ethics Committee of the Faculty of Medicine, Chulalongkorn University (IRB numbers 192/64 and 281/64). This study has been registered with the Thai Clinical Trials Registry (TCTR20210319003 and TCTR20210520004). Informed consent was obtained before participant enrollment.

