## Supplementary Information for "Immune responses to inactivated and vector-based vaccines in individuals previously infected with SARS-CoV-2"

| AEs | CoronaVac | AZD1222 | Risk difference (95%CI) |
|---|---|---|---|
| **Injection site pain** | 32/58 (55.2%) | 50/59 (84.8%) | 29.6 (13.8, 45.3) |
| **Swelling** | 2/58 (3.5%) | 13/59 (22%) | 18.6 (7.0, 30.2) |
| **Redness** | 3/58 (5.2%) | 14/59 (23.7%) | 18.6 (6.3, 30.8) |
| **Fever** | 0/58 (0%) | 10/59 (17%) | 17 (7.4, 26.5) |
| **Headache** | 20/58 (34.5%) | 47/59 (79.7%) | 45.2 (29.2, 61.2) |
| **Myalgia** | 20/58 (34.5%) | 48/59 (81.4%) | 46.9 (31.1, 62.6) |
| **Fatigue** | 1/58 (1.7%) | 9/59 (15.3%) | 13.5 (3.8, 23.3) |
| **Dizziness** | 3/58 (5.2%) | 10/59 (17%) | 11.8 (0.6, 22.9) |
| **Diarrhea** | 3/58 (5.2%) | 5/59 (8.5%) | 3.3 (-5.8, 12.4) |
| **Vomiting** | 1/58 (1.7%) | 2/59 (3.4%) | 1.7 (-4.0, 7.4) |
| **Numbness** | 2/58 (3.5%) | 3/59 (5.1%) | 1.6 (-5.7, 9) |
| **Back pain** | 0/58 (0%) | 3/59 (5.1%) | 5.1 (-0.5, 10.7) |

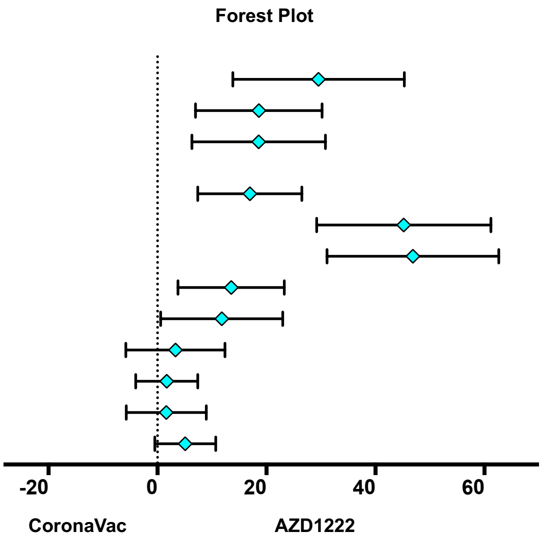


**Supplementary Fig 1. Forest plot showed the risk difference with 95% confidence intervals in the proportion of participants with any grade solicited AEs after first dose vaccination compared with AZD1222 and CoronaVac vaccine.** AE denotes adverse events

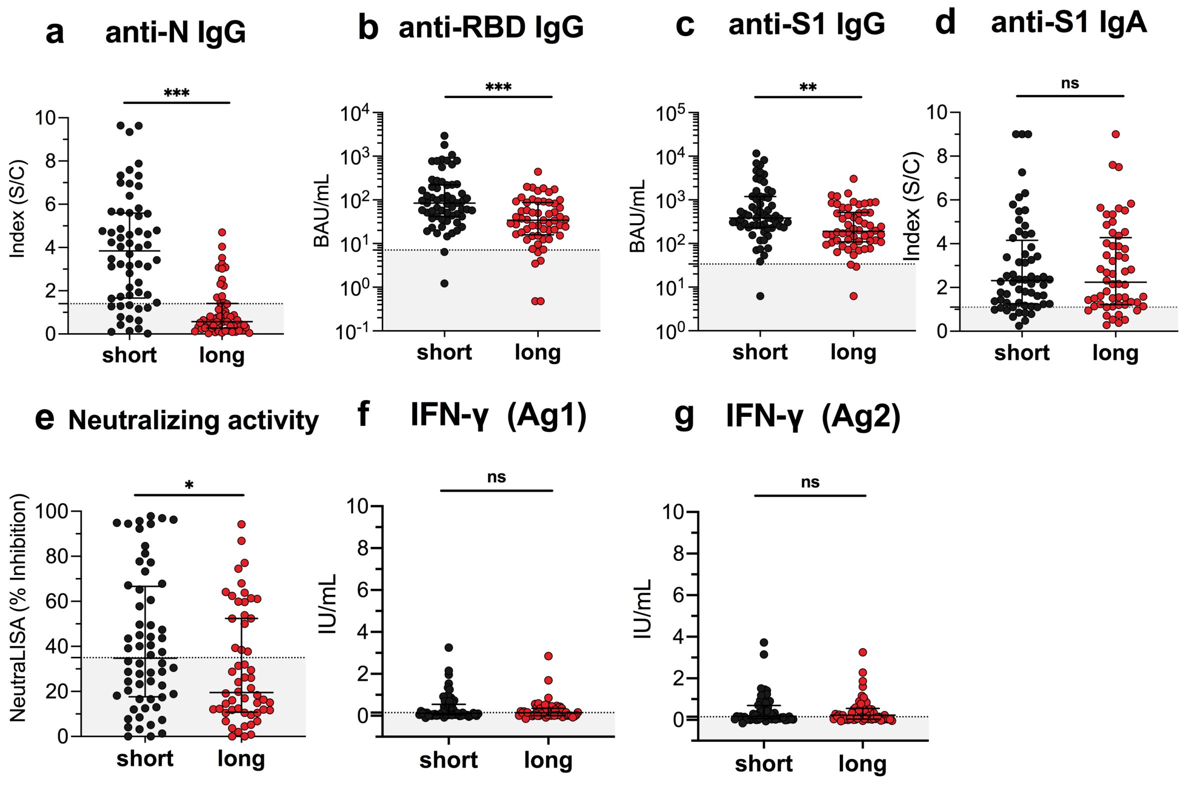


**Supplementary Fig 2. Baseline serological testing in serum was collected from previously infected individuals with short (2-5 months, n=60) and long (13-15 months, n=57) intervals.** Scatter plots display (A) anti-N IgG, (B) anti-RBD IgG, (C) anti-S1 IgG, (D) anti-S1 IgA, (E) neutralizing activity, total interferon-gamma to SARS-CoV-2 (F) Ag1 (stimulated by CD4+ epitopes derived from RBD and (E) Ag2 ( stimulated by CD4+ and CD8+ epitopes from S1 and S2 subunit) as the median value with the interquartile ranges and geometric mean titers (GMT) with 95% confidence intervals. Statistical analysis was analyzed using the Mann-Whitney test.

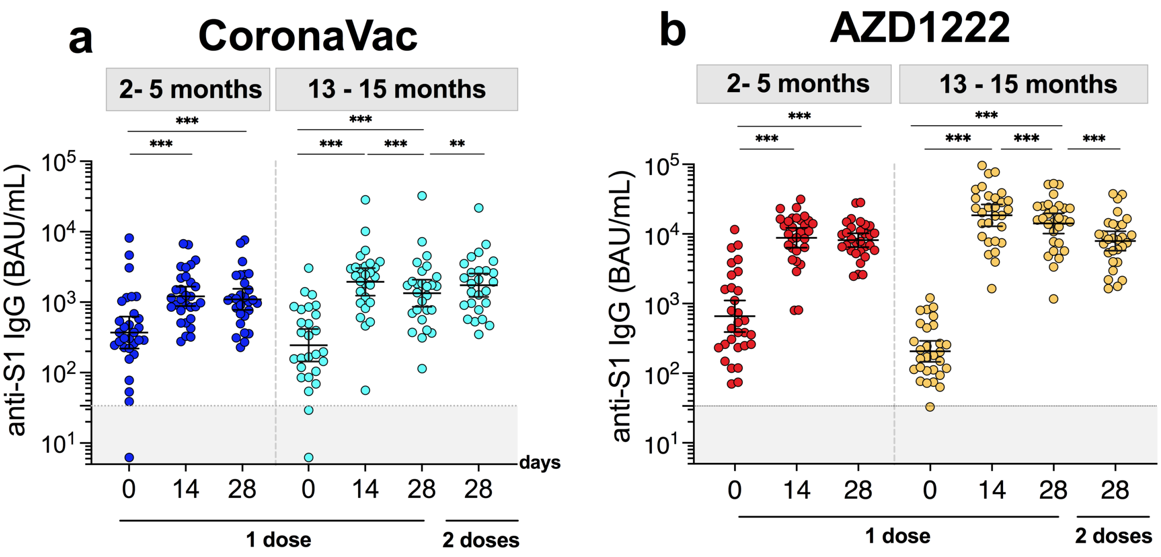


**Supplementary Fig 3. Anti-S1 IgG antibody response to (A) inactivated (CoronaVac) and (B) vector-based (AZD1222) SARS-CoV-2 vaccination in individuals with and prior SARS-CoV-2 infection.** Scatter plots display the geometric mean titers (GMT) with 95% confidence intervals. Statistical analysis was analyzed using the Kruskal-Wallis test with Dunns' post-hoc correction.

**Supplementary Table 1.** GMT (with 95% confidence intervals) of anti-RBD IgG, median (with interquartile ranges) of anti-N IgG and anti-S IgA S/C ratios, percentage inhibition against wild-type and variants of SARS-CoV-2 and interferon gamma responses in serum from vaccinated individuals who received CoronaVac (CV) or AZD1222 with different time point of blood sampling.

| **Vaccine** | **CoronaVac** | **CoronaVac** | **CoronaVac** | **AZD1222** | **AZD1222** | **AZD1222** |
|---|---|---|---|---|---|---|
| **Interval between the date of positive test and the first vaccine dose** | **2-5 months (short -interval) (n=30)** | **13-15 months (long-interval) (n=28)** | **No history of infection (n=90)** | **2-5 months (short -interval) (n=30)** | **13-15 months (long-interval) (n=29)** | **No history of infection (n=90)** |

| | | | | | | |
|---|---|---|---|---|---|---|
| **Female, n (%)** | 15 (50) | 8 (28.6) | 41 (45.6) | 13 (43.3) | 18 (62.1) | 50 (55.6) |
| **Age, mean(range)** | 37.6 (20-56) | 43.1 (25-57) | 42.6 (24-59) | 39.4 (25-66) | 44.7 (27-66) | 47.6 (19-85) |
| **Anti-RBD IgG (BAU/mL)** | | | | | | |
| Day 0 (1st dose) GMT (95% CI) | 75.4 (44-128) | 33 (18-60) | N/A | 120.3 (71-203) | 30.6 (21-45) | N/A |
| n | 30 | 28 | N/A | 30 | 29 | N/A |
| Day 14 (1st dose) GMT (95% CI) | 253 (182-350) | 239 (144-397) | N/A | 1921 (1511-2442) | 2651 (1873-3752) | N/A |
| n | 30 | 28 | N/A | 30 | 29 | N/A |
| Day 28 (1st dose) GMT (95% CI) | 235 (164-336) | 257 (171-385) | N/A | 1549 (1245-1926) | 2258 (1640-3108) | N/A |
| n | 28 | 28 | N/A | 30 | 29 | N/A |
| Day 28 (2nd dose) GMT (95% CI) | N/A | 311 (223-434) | 116 (100-136) | N/A | 1294 (958-1748) | 184 (153-222) |
| n | N/A | 27 | 90 | N/A | 29 | 89 |
| **Anti-trimeric S1 IgG (BAU/mL)** | | | | | | |
| Day 0 (1st dose) GMT (95% CI) | 369.2 (219-622) | 244.6 (144-417) | N/A | 658 (390-1108) | 206 (146-291) | N/A |
| n | 30 | 27 | N/A | 30 | 29 | N/A |
| Day 14 (1st dose) GMT (95% CI) | 1208 (880-1658) | 1942 (1239-3044) | N/A | 8802 (6354-12194) | 18473 (12886-26484) | N/A |
| n | 30 | 27 | N/A | 30 | 29 | N/A |
| Day 28 (1st dose) GMT (95% CI) | 1096 (775-1550) | 1340 (859-2089) | N/A | 8140 (6475-10234) | 14135 (10122-19740) | N/A |
| n | 28 | 27 | N/A | 30 | 29 | N/A |
| Day 28 (2nd dose) GMT (95% CI) | N/A | 1736 (1185-2544) | N/A | N/A | 7911 (5735-10912) | N/A |

| | | | | | | |
|---|---|---|---|---|---|---|
| n | N/A | 27 | N/A | N/A | 29 | N/A |
| **Anti-N IgG (S/C)** | | | | | | |
| Day 0 (1st dose) median (IQR) | 3.5 (1.3-5.7) | 0.6 (0.2-1.5) | N/A | 4.1 (2.1-5.5) | 0.6 (0.3-1.3) | N/A |
| n | 30 | 28 | N/A | 30 | 29 | N/A |
| Day 14 (1st dose) median (IQR) | 3.9 (2.3-5.2) | 3.2 (1.5-4.9) | N/A | 3.2 (1.7-4.6) | 0.6 (0.3-1.4) | N/A |
| n | 30 | 28 | N/A | 30 | 29 | N/A |
| Day 28 (1st dose) median (IQR) | 3.4 (2.1- 4.9) | 3.9 (2.1-6.2) | N/A | 2.8 (1.4-4.4) | 0.6 (0.3-1.3) | N/A |
| n | 28 | 28 | N/A | 30 | 29 | N/A |
| Day 28 (2nd dose) median (IQR) | N/A | 4.5 (3.6-7.0) | 0.8 (0.3-2.1) | N/A | 0.5 (0.2-1.0) | 0.03 (0.02-0.07) |
| n | N/A | 27 | 90 | N/A | 29 | 90 |
| **Anti-S1 IgA (S/C)** | | | | | | |
| Day 0 (1st dose) median (IQR) | 2.3 (1.1-3.9) | 1.6 (1.1-3.2) | N/A | 2.3 (1.4-4.9) | 3.2 (1.4-4.7) | N/A |
| n | 30 | 28 | N/A | 30 | 29 | N/A |
| Day 14 (1st dose) median (IQR) | 3.9 (2.2-6.0) | 7.0 (4.7-9) | N/A | 9 (7.8-9) | 9 (9-9) | N/A |
| n | 30 | 28 | N/A | 30 | 29 | N/A |
| Day 28 (1st dose) median (IQR) | 3.0 (1.7-5.8) | 6.3 (4.3-9) | N/A | 7.6 (6-9) | 9 (9-9) | N/A |
| n | 30 | 28 | N/A | 30 | 29 | N/A |
| Day 28 (2nd dose) median (IQR) | N/A | 7.5 (4.8-9) | 1.0 (0.6-2.4) | N/A | 9 (8.5-9) | 1.2 (0.6-2.1) |
| n | N/A | 27 | 30 | N/A | 29 | 30 |

| | | | | | | |
|---|---|---|---|---|---|---|
| **sVNT—Wild Type.** (Euroimmun, Lubeck, Germany | | | | | | |
| Day 0 (1st dose) median (IQR) | 33 (16.6-58.5) | 21.5 (9.6-53.9) | N/A | 37.6 (18-78.6) | 19.1 (11.8-35.2) | N/A |
| n | 30 | 27 | N/A | 30 | 29 | N/A |
| Day 14 (1st dose) median (IQR) | 82.3 (68.4-96) | 95.8 (76.7-98.6) | N/A | 99.4 (99-99.6) | 99.5 (99.4-99.6) | N/A |
| n | 30 | 27 | N/A | 30 | 29 | N/A |
| Day 28 (1st dose) median (IQR) | 84.6 (52-93.6) | 94.9 (82.6-98) | N/A | 99.4 (98.6-99.6) | 99.6 (99.4-99.6) | N/A |
| n | 28 | 27 | N/A | 30 | 29 | N/A |
| Day 28 (2nd dose) median (IQR) | N/A | 95.5 (78.4-98) | 48.8 (28.9-68.8) | N/A | 99.4 (98.7-99.5) | 77.2 (57-89.7) |
| n | N/A | 26 | 90 | N/A | 29 | 89 |
| **sVNT—Wild Type. (**GenScript, Jiangsu, China**)** | | | | | | |
| Day 28 (1st dose) median (IQR) | 87.6 (72.6-94.5) | 97.3 (93.8-97.8) | N/A | 97.7 (97.1-97.9) | 97.9 (97.8-98) | N/A |
| n | 29 | 27 | N/A | 30 | 30 | N/A |
| Day 28 (2nd dose) median (IQR) | N/A | 97.6 (94.9-97.9) | 72.4 (54.7-78.8) | N/A | 97.7 (97.6-97.8) | 92.1 (73.5-96.6) |
| n | N/A | 27 | 24 | N/A | 30 | 17 |
| **sVNT—B.1.1.7 (alpha). (**GenScript, Jiangsu, China**)** | | | | | | |
| Day 28 (1st dose) median (IQR) | 80.9 (63.1-90.1) | 93.6 (81.4-97) | N/A | 97.2 (95.8-97.5) | 97.8 (97.3-97.9) | N/A |
| n | 29 | 27 | N/A | 30 | 30 | N/A |
| Day 28 (2nd dose) median (IQR) | N/A | 94.9 (89.7-96.4) | 43.1 (34.9-52.6) | N/A | 97.6 (96.6-97.8) | 77.2 (63.1-89.9) |
| n | N/A | 27 | 25 | N/A | 30 | 17 |

| | | | | | | |
|---|---|---|---|---|---|---|
| **sVNT–B.1.351 (beta). (**GenScript, Jiangsu, China**)** | | | | | | |
| Day 28 (1st dose) median (IQR) | 65.7 (47.8-80.7) | 84.5 (69.2-93.7) | N/A | 94.2 (89.7-95.7) | 96.2 (95.4-96.6) | N/A |
| n | 29 | 27 | N/A | 30 | 30 | N/A |
| Day 28 (2nd dose) median (IQR) | N/A | 88.6 (72.7-93.4) | 35.5 (25.8-49.7) | N/A | 96.4 (94.7-97) | 69.3 (45.7-86) |
| n | N/A | 27 | 25 | N/A | 30 | 17 |
| **sVNT–B.1.617.2 (delta) (**GenScript, Jiangsu, China**)** | | | | | | |
| Day 28 (1st dose) median (IQR) | 85.4 (66.4-92.2) | 95.3 (85.8-97.4) | N/A | 97.6 (96.3-97.8) | 97.8 (97.6-97.9) | N/A |
| n | 29 | 27 | N/A | 30 | 30 | N/A |
| Day 28 (2nd dose) median (IQR) | N/A | 96.5 (93.4-97.3) | 48.4 (38.9-62.2) | N/A | 97.8 (97.7-98) | 87.7 (64.6-95.3) |
| n | N/A | 27 | 25 | N/A | 30 | 17 |
| **IFN-γ Ag 1** | | | | | | |
| Day 0 (1st dose) median (IQR) | 0.1 (0.02-0.45) | 0.2 (0.02-0.46) | N/A | 0.12 (0.07-0.65) | 0.12 (0.01-0.3) | N/A |
| n | 30 | 27 | N/A | 30 | 29 | N/A |
| Day 14 (1st dose) median (IQR) | 0.34 (0.13-0.95) | 0.76 (0.15-2) | N/A | 1.21 (0.47-2.38) | 0.84 (0.43-2.4) | N/A |
| n | 30 | 27 | N/A | 30 | 29 | N/A |
| Day 28 (1st dose) median (IQR) | 0.31 (0.07-0.79) | 0.42 (0.13-1.5) | N/A | 1.16 (0.41-2.13) | 0.83 (0.28-1.17) | N/A |
| n | 28 | 27 | N/A | 30 | 29 | N/A |
| Day 28 (2nd dose median (IQR) | N/A | 1.24 (0.7-2.1) | N/A | N/A | | N/A |
| n | N/A | | N/A | N/A | | N/A |

| | | | | | | |
|---|---|---|---|---|---|---|
| **IFN- γ Ag 2** | | | | | | |
| Day 0 (1st dose) median (IQR) | 0.17 (0.03-0.83) | 0.25 (0.05-0.77) | N/A | 0.21 (0.07-0.61) | 0.16 (0.03-0.5) | N/A |
| n | 30 | 27 | N/A | 30 | 29 | N/A |
| Day 14 (1st dose) median (IQR) | 0.63 (0.16-1.9) | 1.05 (0.18-3.06) | N/A | 1.99 (0.67-3.26) | 1.3 (0.64-4.42) | N/A |
| n | 30 | 27 | N/A | 30 | 29 | N/A |
| Day 28 (1st dose) median (IQR) | 0.63 (0.14-1.21) | 0.42 (0.08-1.78) | N/A | 1.86 (0.58-3.46) | 1.13 (0.66-2.16) | N/A |
| n | 28 | 27 | N/A | 30 | 29 | N/A |
| Day 28 (2nd dose) median (IQR) | N/A | | N/A | N/A | | N/A |
| n | N/A | | N/A | N/A | | N/A |

N/A= No data availble

**Supplementary Table 2.** Total interferon gamma responses in serum from vaccinated individuals who received CoronaVac (CV) or AZD1222 (AZ) after 28 days of first and second dose. Individuals with elevated the interferon-gamma responses in first dose vaccination were then selected to determine the interferon-gamma responses after second dose vaccination.

| Vaccine | CoronaVac | AZD1222 |
|---|---|---|
| **Interval between the date of positive test and the first vaccine dose** | **13-15 months (long-interval)** | **13-15 months (long-interval)** |

| | | |
|---|---|---|
| **IFN-γ Ag 1** | | |
| Day 28 (1st dose) median (IQR) | 1.52 (0.8-2.1) | 0.95 (0.4-1.2) |
| n | 12 | 27 |
| Day 28 (2nd dose median (IQR) | 1.24 (0.8-2.1) | 0.59 (0.2-1.1) |
| n | 12 | 27 |
| **IFN- γ Ag 2** | | |
| Day 28 (1st dose) median (IQR) | 1.86 (0.9-2.8) | 1.22 (0.8-2.2) |
| n | 12 | 27 |
| Day 28 (2nd dose) median (IQR) | 2.38 (1.3-4.1) | 0.86 (0.3-1.74) |
| n | 12 | 27 |
